# A systematic review and meta-analysis of longitudinal cohort studies comparing mental health before versus during the COVID-19 pandemic

**DOI:** 10.1101/2021.03.04.21252921

**Authors:** Eric Robinson, Angelina R. Sutin, Michael Daly, Andrew Jones

## Abstract

**Background:** Increases in mental health problems have been observed in some studies during the COVID-19 pandemic. It is unclear whether changes have been large and experienced by most population sub-groups, persisted over time or been symptom specific.

**Methods:** We systematically reviewed and meta-analysed longitudinal cohort studies that examined changes in mental health among the same group of participants before and during the pandemic (PROSPERO: CRD42021231256). Searches for published and unpublished studies were conducted in January 2021. Changes in mental health (standardised mean change; SMC) were examined using meta-analyses.

**Findings:** Sixty-five studies were included. There was an overall increase in mental health symptoms that was most pronounced during March-April 2020 (SMC = .102 [95% CI: .026 to .192], p = 0.03) before significantly declining over time (May-July SMC = .067 [95% CI: -.022 to .157], p = .141). Compared to measures of anxiety (SMC = 0.13, p = 0.02) and general mental health (SMC = -.03, p = 0.65), increases in depression and mood disorder symptoms tended to be larger (SMC = 0.22, p < .001) and reductions over time appeared less pronounced. Increased mental health symptoms were observed across most population subgroups examined but there was no evidence of any change in symptoms among samples with a pre-existing mental health condition.

**Interpretation:** There was a small increase in mental health symptoms soon after the outbreak of the COVID-19 pandemic that decreased and was comparable to pre-pandemic levels by mid-2020 among most population sub-groups and symptom types.

**Funding:** N/A

**Research in context:** *Evidence before this study:* There have been reported increases in mental health problems during the outbreak of the COVID-19 pandemic. However, it is unclear whether changes in mental health problems have been symptom specific, how changes have differed across populations and whether increased mental health problems have persisted over time. We systematically reviewed and meta-analysed longitudinal cohort studies that examined mental health among the same participants prior to and during the pandemic in 2020. This approach allowed us to quantify the mental health burden associated with the outbreak of the pandemic and how it has changed over time. We searched Pubmed, SCOPUS, Web of Science and PsychInfo from January 2020 to January 11, 2021 and identified eligible unpublished articles available on pre-print servers.

*Added value of this study:* We identified 65 eligible articles that reported 201 comparisons of mental health pre vs. post pandemic outbreak. Meta-analysis indicated that longitudinal cohort studies that examined mental health prior to and during the COVID-19 pandemic in 2020 showed a significant but statistically small increase in mental health symptoms. The overall increase in mental health symptoms was most pronounced during the early stages of the pandemic (March-April), before decreasing and being generally comparable to pre-pandemic levels by mid-2020. Compared to anxiety and general measures of mental health functioning, increases tended to be larger in depressive symptoms and although statistically small, remained elevated past the early stages of the pandemic. Increases in mental health symptoms were observed across most population sub-groups, but there was no evidence of a change in mental health symptoms among samples of participants with a pre-existing mental health condition.

*Implications of all the available evidence:* Findings confirm that the initial outbreak of the pandemic was associated with a significant but statistically small increase in mental health symptoms. Given that small effects may have meaningful cumulative consequences at the population level, there is a need for continued mental health provision and monitoring particularly during periods of the pandemic when infection rates and deaths are high. Further into the pandemic, mental health problems decreased significantly, which indicated recovery and resilience in overall mental health. Contrary to predictions made early in the pandemic, there was also no evidence of a worsening of mental health symptoms among samples of participants with a pre-existing mental health condition. Overall the results of the present analyses suggest that the pandemic may not have caused an unprecedented and long lasting mental health crisis, instead there appears to have been resilience in mental health.

## Introduction

As of February 2020, the COVID-19 pandemic has been responsible for more than 2.5 million deaths worldwide (1). Soon after the COVID-19 crisis was declared a pandemic by the World Health Organisation (WHO) on the 11^th^ March, concerns were raised over a potential parallel mental health crisis fuelled by the pandemic and the associated social restrictions imposed by governments to reduce virus transmission (2, 3). The mental health of ‘at risk’ groups during the pandemic, such as those with pre-existing mental health conditions, has also been highlighted as cause for concern (3, 4).

There are considerable methodological challenges to quantifying the impact that the pandemic has had on mental health. During the outbreak of the pandemic studies indicated that many participants perceived their mental health had worsened (5, 6), but retrospective reports of change in mental health are prone to substantial bias (7, 8). Other studies have found a greater incidence of mental health problems in cohorts recruited during the pandemic compared to different cohorts who completed measures prior to the pandemic (9, 10). However, differences in how samples were recruited (e.g. greater reliance of online and non-probability based samples during the pandemic) and differences in demographic profiles of pre vs. post-pandemic outbreak cohorts also make inferring change in mental health attributable to the pandemic difficult (11).

A number of longitudinal cohort studies have sampled the same participants before and during the pandemic to examine how mental health has changed. In a large nationally representative sample of UK adults, Daly et al. found that non-specific general mental health symptoms (GHQ-12) increased in April-June 2020 compared to a pre-pandemic baseline (12). Other longitudinal cohort studies have found little change in mental health (13) or mixed evidence (14). Longitudinal studies are uniquely placed to characterise the time course of the mental health burden associated with the COVID-19 pandemic. Although the effects of the pandemic on mental health could be long lasting (15), a recent multi-wave longitudinal study of US adults found that after an initial increase in distress during the early stages of the pandemic, distress reduced to pre-pandemic levels within a few months (16).

There has been no systematic review and meta-analysis of longitudinal cohort studies with pre-pandemic measurement of mental health (17-19). We systematically reviewed and meta-analysed studies that examined longitudinal changes in mental health among the same sample of participants before vs. during the pandemic, in order to quantify the size of the mental health burden associated with the pandemic and how this burden changed over time. Secondary aims were to examine whether changes have been symptom specific and whether changes differed based on population demographics.

## Methods

### Eligibility Criteria

#### Participants

To be eligible, studies were required to have sampled the same cohort of participants prior to 11/03/20 (date the WHO declared a pandemic) (20) and at least once after this date. Chinese studies were eligible (but analysed separately) if mental health was assessed prior to and after 23/01/20 because substantial social restriction measures were enforced across China from this point (21). There were no limits on populations sampled.

#### Measures of interest

To be eligible, studies were required to have collected data using a validated multi-item measure of mental health symptoms or mental well-being, such as depression (e.g. Patient Health Questionnaire: PHQ9), anxiety (Depression, Anxiety, Stress Scale: DASS), non-specific general mental health related functioning and distress (General Health Questionnaire: GHQ12, Kessler) and well-being (Warwick-Edinburgh Mental Wellbeing Scale). As our focus was on mental health symptoms, ineligible measures included loneliness, stress and physical health related quality of life.

#### Outcome

Studies that examined continuous changes (i.e. standardised mean change; SMC) in mental health symptoms were eligible, as was change in the % of the sample meeting questionnaire specific cut-offs for clinically relevant/ likely serious mental health problems were eligible (i.e. Odds Ratio).

#### Study design features

Studies were required to sample the same participants using the same measure of mental health pre and post-pandemic (repeated cross-sectional studies were not eligible). If only a sub-sample of participants were followed up across survey waves, only data from the sub-sample were eligible. Interventions to improve mental health during the pandemic were not eligible. If multiple articles reported on data from the same cohort of participants, the article with the largest number of post-pandemic follow-up data collection points was included. Journal articles and pre-prints were eligible.

### Article Identification

We searched Pubmed, SCOPUS, Web of Science and PsychInfo from January 2020 to January 11, 2021, using combinations of coronavirus and mental health relevant search terms (see online supplementary materials). One author conducted title and abstract screening and 25% were cross-checked by a second author (no discrepancies). Two authors conducted full-text screening and disagreements in eligibility were resolved by discussion. We searched three databases for unpublished pre-prints; Open Science Framework (inclusive of 30 preprint archives, e.g. PsychArxiv), MedrXiv and the Social Science Research Network, and conducted forward citation tracking (Google Scholar) for all eligible articles. A single author conducted pre-print searches and forward citation tracking; a second author verified eligibility of identified articles.

### Data Extraction

The following information was extracted by a single author and checked for accuracy by a second author; country of study, participant group sampled, age group of sample, sampling strategy used (e.g. use of representative sampling vs. convenience), pre and post pandemic dates of data collection, mental health measure, analytic treatment of mental health symptomology change (e.g. use of change in % meeting questionnaire cut-off vs. continuous change in questionnaire score), level of attrition (%), effect size information (authors were contacted if information was missing) and whether the study was reported in a journal article or pre-print.

### Risk of Bias

We reviewed methodological quality scales and risk of bias measures to develop a list of bias indicators relevant to included studies. See online supplementary materials for full detail (including justifications for each indicator). Indicators were rated by two authors; *i)* whether the study reported representative sampling (yes = lower in risk of bias), *ii)* whether the study underwent peer review (yes = lower in risk of bias), *iii)* low level of attrition unlikely to affect results (yes = lower in risk of bias), *iv)* whether the study had a relatively large (N≥1000) sample size (yes = lower in risk of bias*), v)* whether the pre-pandemic measure of mental health was collected within the last year of the post-pandemic outbreak measure (yes = lower in risk of bias), *vi)*, whether survey delivery mode (e.g. online) was consistent across pre and post outbreak waves of data collection (yes = lower in risk of bias), *vii)* whether conflicts of interest were reported (no conflicts = lower in risk of bias).

### National COVID-19 data

To examine if heterogeneity of change in mental health was attributable to country level factors, for each eligible study, we identified the number of recorded COVID cases and deaths (by country) per million population for the month that mental health was measured during the pandemic. We also used data from the Oxford COVID-19 Government Response Tracker to characterise each country’s severity of social restrictions, number of health measures and level of economic support in place during the month post-pandemic outbreak mental health was assessed. See supplementary materials for full information.

### Analyses

For our main analysis on continuous data, we computed standardised mean change (SMC) in pooled SD units For analyses examining questionnaire cut-off data we computed Marginal Odds Ratios to quantify size of change. See online supplementary materials for full statistical information. As studies contributed multiple comparisons to analyses, we conducted a multi-level meta-analysis. Heterogeneity was assessed using I^2^ statistic. We examined whether time (month post-pandemic outbreak measure was collected) predicted change in mental health symptoms using meta-regression. We also conducted sub-group analyses on symptom type (depression, anxiety, general mental health functioning (including distress), well-being, psychotic symptoms, other), gender, age (adult vs. child/adolescent), population sampled (general population, university students, mental and physical health conditions) and continent sample was from. A minimum of n=5 effects for each sub-group was required for analysis. All analyses were conducted in R. To address influential cases we conducted leave one-out-analyses and computed DFBETAS values for each effect size in the full models (without moderators). Funnel plots were inspected for potential publication bias, Egger’s test of asymmetry and a Trim and Fill procedure were used. See online supplementary materials for full details.

## Results

### Article identification

After removal of duplicates, title and abstract screening of electronic database search results and identification of eligible articles through other sources, 153 articles were full-text screened. A total of 65 articles were eligible for inclusion. See Figure 1 for study selection flow chart.

**Figure 1:**
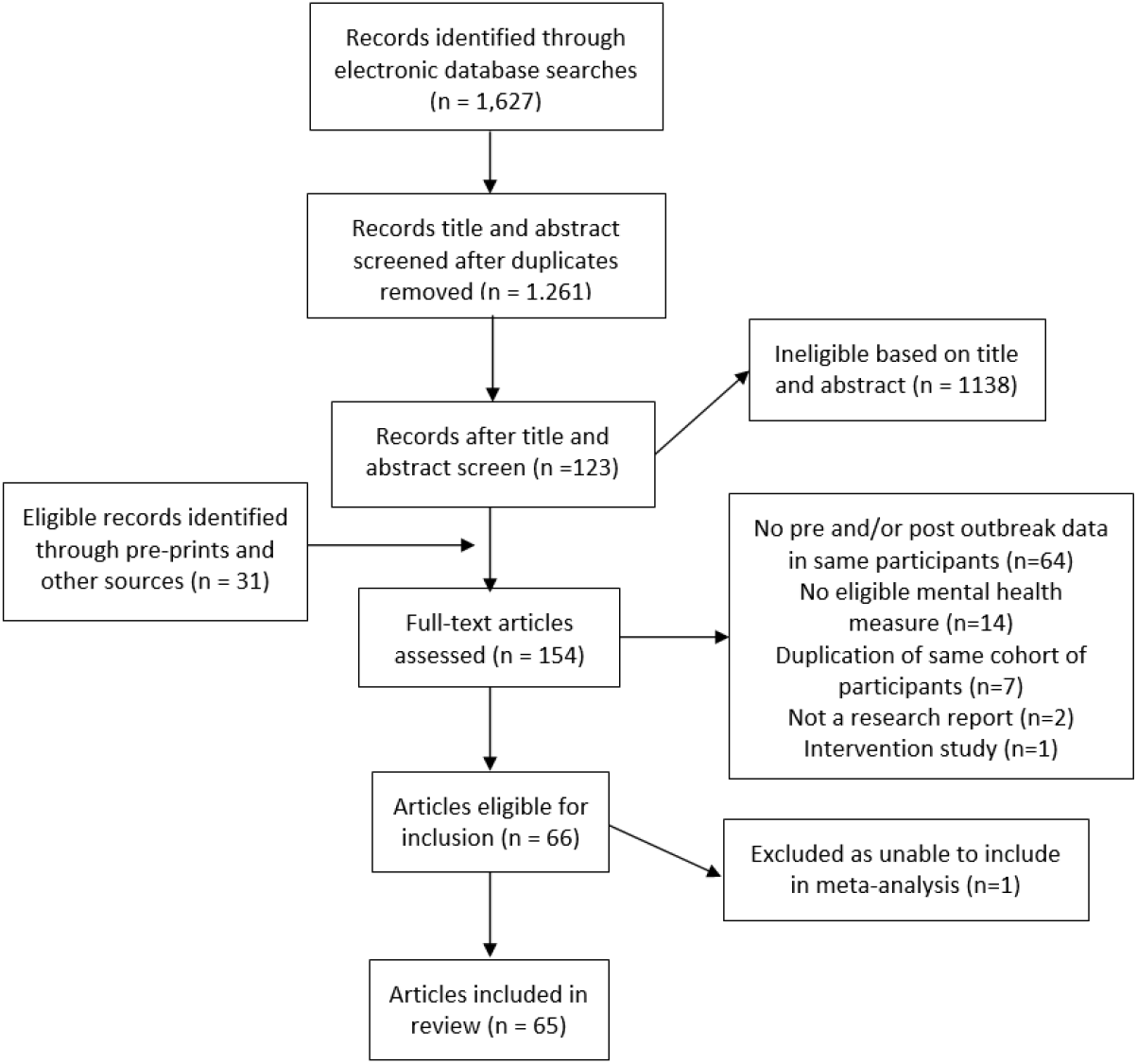
Study search and inclusion flow chart. Not a research report indicates manuscript did not report on a study. Excluded as unable to include in meta-analysis (N=1) was due to authors not providing the required statistical information to compute effect size.

### Overview of studies and eligible effects

The 65 articles reported on a total of 201 pre vs. during pandemic mental health comparisons to include in the meta-analyses. The majority of studies sampled European (N=31) or North American (N=16) populations. The majority of comparisons (n=177) examined pre vs. post pandemic outbreak change in overall mental health as a continuous variable (i.e. change in mean questionnaire score) and n=24 examined change in % of sample meeting questionnaire cut-offs for clinically relevant symptomology. See Table S1 in the supplementary materials for individual study information.

### Overall change in mental health symptoms (SMC)

There were 177 pre vs. post pandemic comparisons across 61 studies. Twelve comparisons (4 studies) came from Chinese samples, which left a total of 165 comparisons included in the main analysis. Sample sizes of comparisons ranged from n=9 to 11,599. Depression and mood disorder symptoms (n=58), anxiety disorder symptoms (n=52) and general non-specific mental health symptoms (n=35) were the most common symptoms studied. From the 165 comparisons drawn from ∼55,015 participants, overall change in mental health symptoms from pre-post pandemic outbreak was significant (SMC = .106 [95% CI: .039 to .172], z = 3.12, p = .002, I^2^ = 97.8) and indicative of heterogeneous and small increase in symptoms (SMC=0.2 is indicative of a small effect). There were no influential cases, and although there was some evidence of funnel plot asymmetry, Trim and Fill did not impute any studies (see online supplementary materials).

For studies in China (∼ 1,854 participants), change in mental health symptoms was indicative of a small, non-significant increase in symptoms (SMC = .194 [95% CI: -.576 to .964], z = 0.49, p = .622].

### Symptom-level analyses

We examined whether SMC in mental health symptoms was moderated by symptom type. The test of moderation was significant (QM(5) = 19.71, p < .001). There was a significant increase in symptoms of anxiety (SMC = .125 [95% CI: .019 to .230], z = 2.31, p = .021) and depression (SMC = .216 [95% CI:.135 to.296], z = 5.24, p < .001), with the increase in depression larger than anxiety (p = .049). There was a significant decrease in symptoms of psychosis (SMC = -.211 [95% CI: -.376 to -.046], z = 2.51, p = .012).^1^ There was no significant change in measures of general mental health (SMD = -.030 [95%CI -.158 to .098], z = 0.457, p = .648), well-being (SMC = .067 [95% CI: -.123 to .256],z = .690, p = .490) or for the mixed ‘other’ conditions sub-group (SMC = -.041 [95% CI: - .203 to .121], z = 0.501, p = .616). See online supplementary materials figures S2-S7 for symptom level forest plots.

### Time analyses

Change in symptoms from pre-pandemic levels became smaller over each month (monthly change coefficient = -.057 [95%CI: -.100 to -.013], z = 2.57, p = .010). To illustrate, among post-pandemic measures of mental health collected in March and April (n = 98) the change in mental health was statistically small and significant; SMC = .102 [95% CI: .026 to.192] z = 2.22, p = .026). Conversely, for measures collected during May-July (n = 67) there was no significant change compared to pre-pandemic mental health symptoms; SMC = .067 [95% CI: -.022 to .157], z = 1.47, p = .141). There was no robust interaction between symptom types and time. See Table 1 for SMCs for each symptom type by time.

**Table 1.**
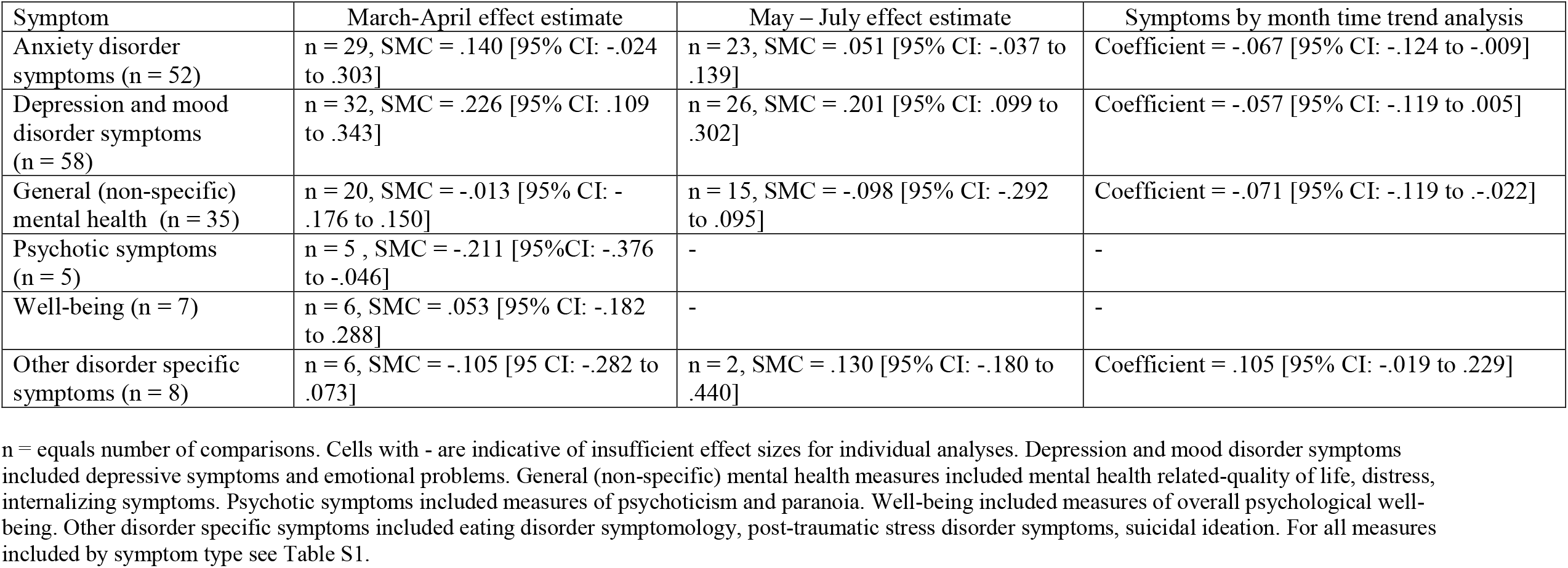
Standardised mean change in symptoms when mental health was measured earlier vs. later in the pandemic.

### Sub-group analyses

Across sub-groups, we found no evidence that change in mental health symptoms differed based on age, gender, or study continent. See Table 2. Change in symptoms tended to be larger among participants with a pre-existing physical health condition compared to the general population. Notably, change in mental health symptoms was non-significant in samples with pre-existing mental health conditions. No country-level data (number of COVID cases/deaths, stringency of government measures or level) explained heterogeneity between samples (ps >.05). See online supplementary materials for full results.

**Table 2.**
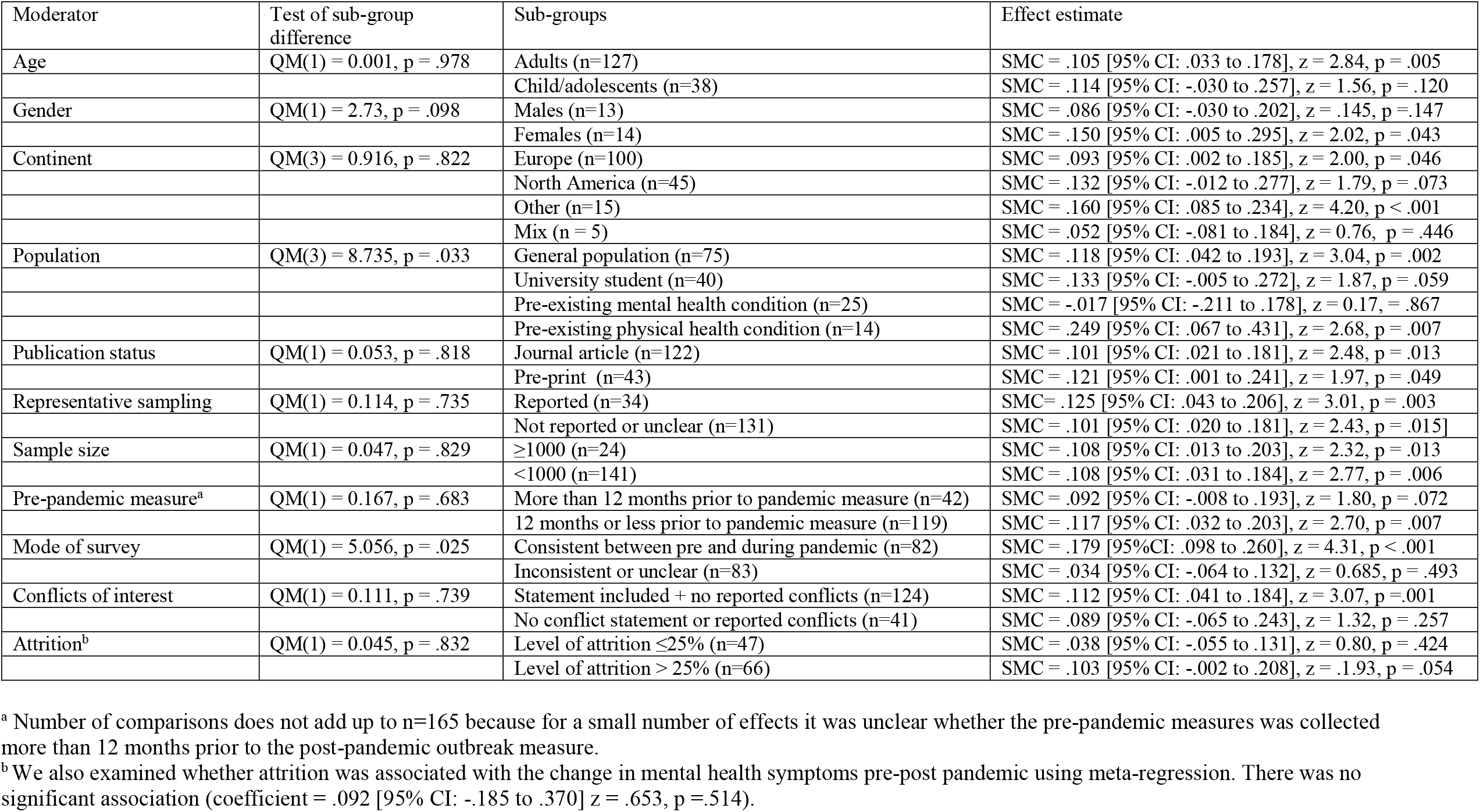
Sub-group analyses for standardised mean change (SMC) in mental health symptoms.

### Risk of bias

Due to the small number of studies examining changes in % of sample meeting questionnaire cut-offs, risk of bias analyses were limited to SMC studies. See Table 2 for risk of bias summary information and results and Table S2 for study-level information. We found limited evidence that the risk of bias indicators predicted size of change in symptoms, except that change in mental health tended to be larger when delivery mode of questionnaire was consistent pre vs. post pandemic. In an exploratory analysis limited to a more homogenous collection of studies that were of lower risk of bias results were consistent with the main analyses. See Figure 2.

**Figure 2:**
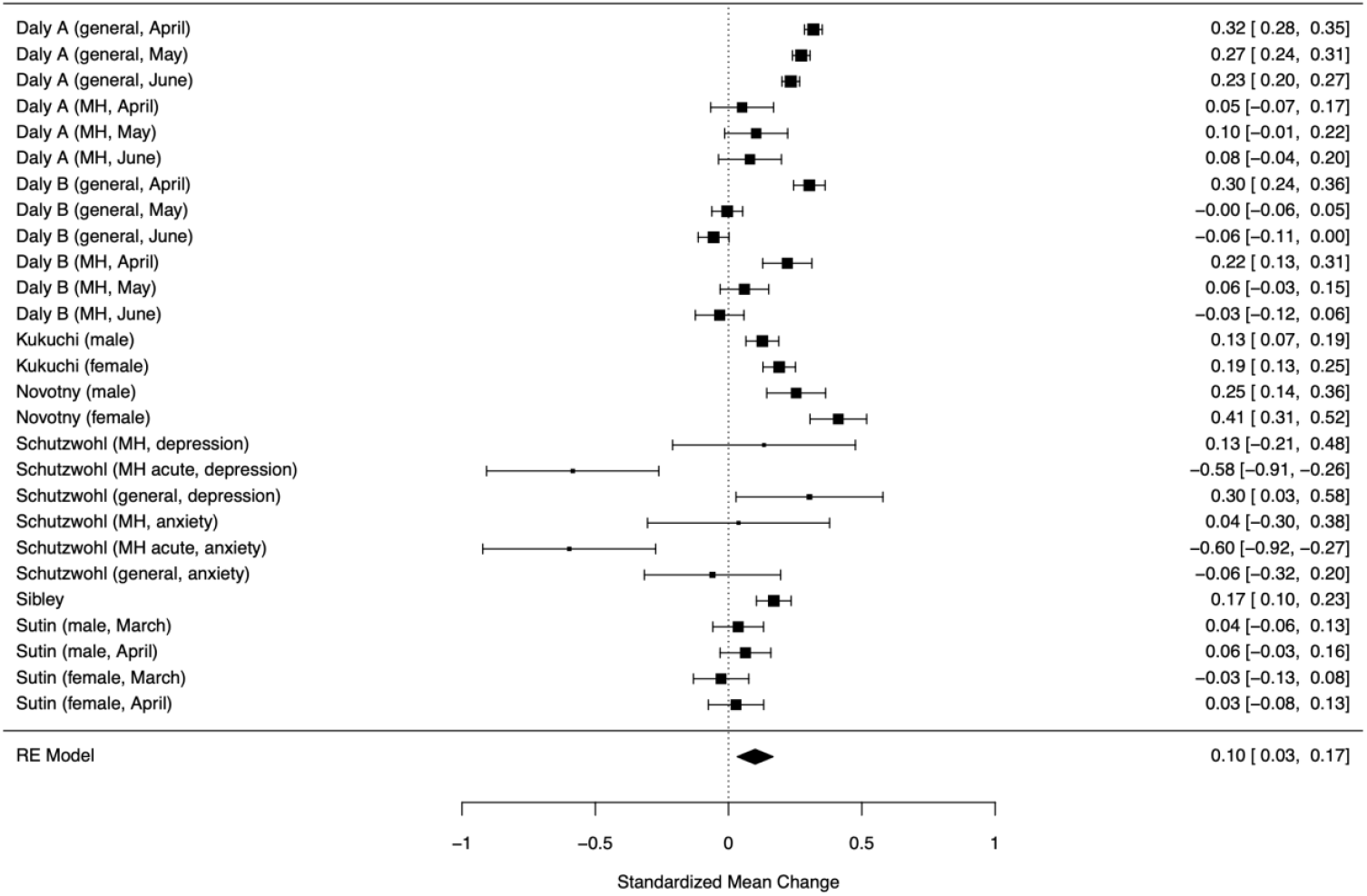
Forest plot of effect sizes from studies of depression, general mental health and anxiety symptoms with lower risk of bias. Analyses of more homogenous collection of studies (depression, anxiety, general mental health measures only) that were of lower risk of bias (used representative sampling, did not report inconsistent mode of survey delivery, reported no conflicts of interest) of ∼27,736 participants, SMC = .100 [95% CI: .033 to.166], z = 2.95, p = .003, I2 =98.0%). General (general population sample), MH (sample with pre-existing mental health condition).

### Change in numbers exceeding questionnaire cut-offs for mental health problems

Twenty-four comparisons across 12 studies (∼21,825 participants) were included. There was a significant effect (single level meta-analysis), with increased odds of exceeding a questionnaire cut-off for mental health problems from pre-post pandemic (Marginal Odds Ratio = 1.31 [95% CI: 1.10 to 1.55], z = 3.18, p = .001, I^2^ = 93.2%), where an OR of 1.5 is tconsidered a small sized effect. See Figure 3. There was no evidence of publication bias or influential cases. See online supplementary materials for full results.

**Figure 3:**
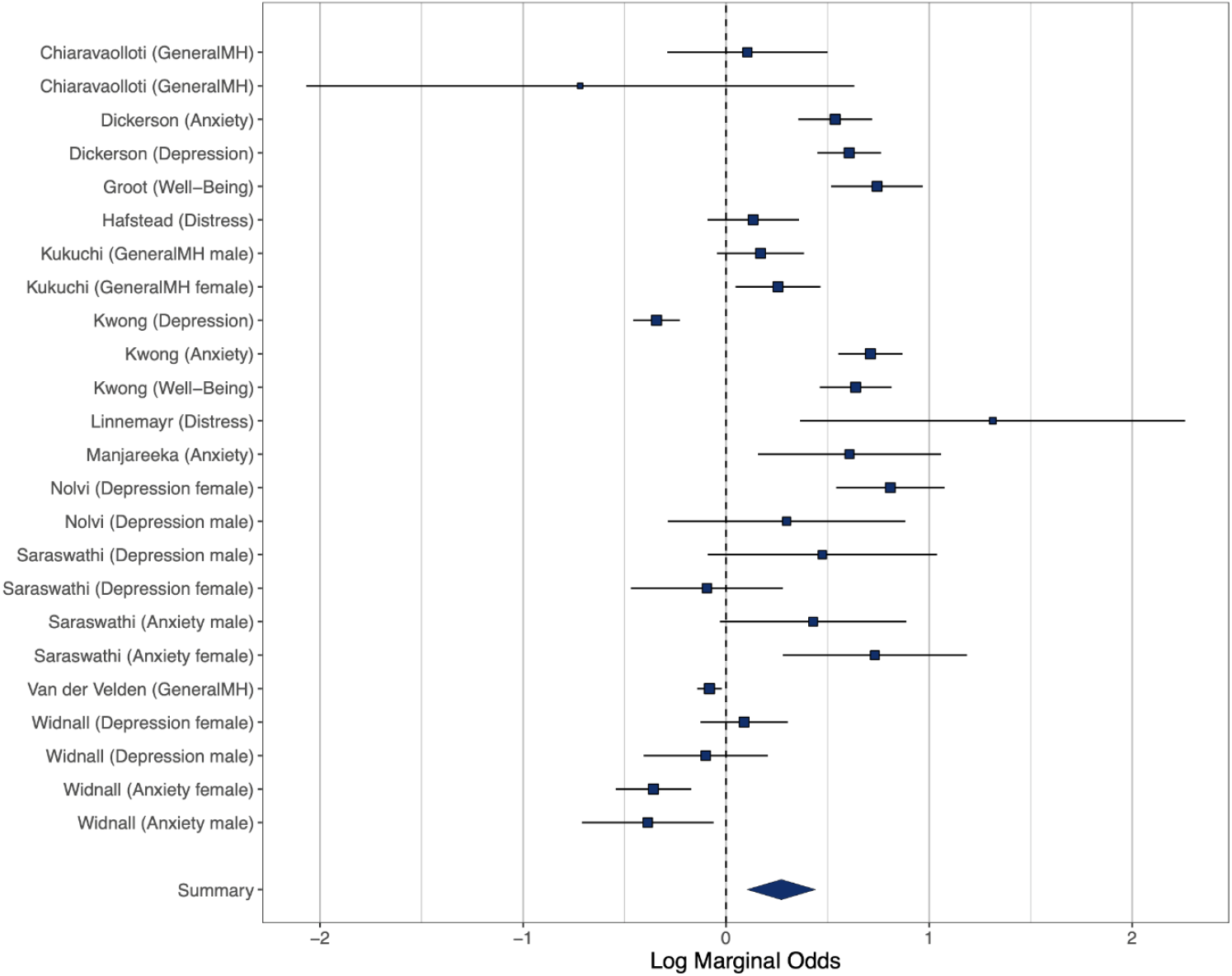
Forest plot of effect sizes for exceeding questionnaire cut offs. General MH (measure of general mental health functioning)

## General Discussion

Across sixty-five longitudinal cohort studies comparing mental health prior to and during the COVID-19 pandemic there was a statistically small overall increase in mental health symptoms. The increase in mental health symptoms was largest among studies that sampled participants in the early stages of the pandemic (March-April) and severity of mental health symptoms decreased significantly over the following months (May-July). This pattern of results may represent an acute and normal response to an unforeseen and distressing traumatic event (22), which was then followed by a period of psychological adaptation and resilience (16, 23). In line with this interpretation, a large study of US adults found that perceived risk and worries about financial instability, infection and death were highest during the early stages of the pandemic(24). However, as more information about the pandemic became available perceived risks decreased and this predicted recovery to pre-pandemic levels of distress (24). Similarly, in a large sample of UK adults recruited after the pandemic outbreak, both anxiety and depressive symptoms showed a trajectory of recovery from the beginning of April onwards (25).

Worsening of mental health symptoms was largest in studies examining depression and mood disorders symptoms, and a small increase in depression symptoms was still observed during May-June. Change in anxiety symptoms was smaller and was not significant in samples measured in May-June. Changes in non-specific general mental health functioning (including distress) and well-being were small and non-significant. The more pronounced change in depressive symptoms observed may reflect the effects of isolation and sadness caused by social restrictions and loss of life during the pandemic (26). Increases in mental health problems were observed across most population sub-groups sampled (e.g. general population, university students, existing physical health condition).

Contrary to concerns raised early in the pandemic (3, 4), changes in mental health were less pronounced among people with pre-existing mental health conditions and overall there was no statistically significant change in mental symptoms in this group. These findings may in part be explained by regression to the mean and naturally occurring recovery of more severe mental health symptoms over time (27), as well as stay at home restrictions that potentially provided a more structured routine and reduced exposure to external stressors (e.g. large social gatherings) among those with severe mental health conditions (22). A number of countries, such as China, introduced new mental health services in response to the pandemic (4) and this may have relieved symptoms among those with and without pre-existing mental health conditions. In contrast, mental health symptoms increased among those with pre-existing physical health conditions, which may reflect the elevated risk posed by COVID-19 to this group.

Increases in mental health symptoms were observed in both North American and European samples, though there were a limited number of studies from other continents. We examined whether a range of country-level factors explained heterogeneity in mental health change across samples, including number of country level COVID-19 cases and attributed deaths in the month that mental health was measured, levels of government financial support and social restrictions in place to reduce virus transmission. We found no evidence that country level factors explained heterogeneity. However, it is difficult to make firm conclusions because the majority of studies were conducted in the early phase of the pandemic when deaths from COVID-19 were high and restrictive measures had already introduced relatively high levels of restriction. A number of countries also implemented restrictions regionally.

A smaller sub-set of studies examined change in proportion of sampled population exceeding questionnaire cut-offs for clinically relevant mental health symptoms. In line with the main analyses, there was a statistically small increase in likelihood of meeting questionnaire cut-offs. A limitation of these studies is that although questionnaires used have been shown to be valid indicators of clinically relevant mental health disorder symptomology, it was common for response formats to be altered to ask participants to report on shorter time frames (e.g. the last week). Therefore, these studies provide an indication of acute symptom severity rather than clinical diagnostic value (e.g. anxiety disorder diagnosis typically requires symptoms for several months (28)).

In contrast to the fear that the COVID-19 pandemic would cause a parallel and longstanding mental health crisis (2, 3), the present findings suggest that overall there has been considerable resilience in mental health. Data on recorded suicides align with this, as there have been stable rates or decreases reported across a number of countries (29). However, there is a need for continued mental health provision and monitoring particularly during periods of increased COVID-19 infection and death. The increase in depression and mood disorder symptoms that did not return to pre-pandemic levels warrants attention, as even a small upward shift in depressive symptoms may have meaningful cumulative consequences on the population-level.

There are limitations to the studies included. The majority of studies sampled populations in developed countries during the early stages of the pandemic. Given that there have been second waves of the pandemic in many countries during early 2021, it will be important to continue to monitor mental health. Heterogeneity of models ended to be high and although some of this variability was explained by symptom type and population groups sampled, it will be important to identify further explanatory factors. For example, there are population sub-groups who may be at greater risk of declines in mental health that we were unable to examine and who are likely to be under-represented in studies of the general population. Due to stressful working conditions frontline healthcare workers may be at increased risk and there may be long-term mental health effects among those who become seriously ill from COVID-19, live in nursing homes or those who suffered financially as a result of the pandemic (30). Participants with pre-existing mental health conditions tended to be grouped in studies and therefore we were unable to examine changes in symptoms among specific patient groups (e.g. schizophrenia). We found little evidence that study outcomes were strongly related to individual risk of bias indicators and in an analysis limited to relatively high-quality studies results were similar to the main analyses. Level of attrition was high across studies and this is a limitation. Although there was no significant evidence of attrition being associated with change in symptoms in meta-regression or sub-group analyses and lower levels of attrition tended to be associated with a smaller as opposed to larger increase in symptoms.

## Conclusions

Among longitudinal cohort studies examining mental health prior to and during the COVID-19 pandemic in 2020 there was a significant but statistically small increase in mental health symptoms. Increases were larger for depressive symptoms compared to anxiety and measures of overall mental health functioning. The overall increase in mental health symptoms was most pronounced during the first two months of the pandemic, before decreasing and being comparable to pre-pandemic levels by mid-2020. There was no evidence of a worsening of mental health symptoms among samples of participants with a pre-existing mental health condition.

## Data Availability

Study data file and pre-registered analysis protocol are available online https://osf.io/rg24v/. Analysis code and files are available on request.

## Contributors

The study was designed by all authors. AJ analysed the data. The first draft was written by ER. All authors critically revised the manuscript and agree to be accountable for all aspects of the work.

## Footnotes

1 Multi-level models remained a better fit of the data separately for anxiety, depression and general mental health measures, but not for psychosis, well-being or other symptoms which is likely due to smaller number of effect sizes. Regardless, treating the data as single or multilevel in these cases did not substantially influence the effect sizes or statistical significance of the models.

