## Supplementary material for "A systematic review and meta-analysis of longitudinal cohort studies comparing mental health before versus during the COVID-19 pandemic": Online supplementary materials

### Online Supplementary Material

#### Example combination of search terms

(COVID* OR coronavirus OR SARS-COV-2) AND (Longitudinal OR prospective) AND (Mental health OR mental illness OR mental disorder OR depression OR anxiety OR emotion OR well-being OR distress OR suicide* OR PTSD OR trauma).

#### Risk of bias indicators

We reviewed methodological quality scales and risk of bias measures (1-3) to develop a list of bias indicators relevant to included studies:

**1)** Was the sample recruited to be representative of the population being studied (e.g. use of quota or probability sampling to reflect demographics of wider population) or was the sample likely non-representative (e.g. use of convenience and snowball sampling)? Studies with non-representative sampling (convenience, snowball) were considered higher in risk of bias.

**2)** Was the study reported in a journal article or pre-print? Pre-print articles were considered higher in risk of bias as study methods have not yet been subject to formal peer review.

**3)** Is attrition likely to have substantially influenced study results? E.g. confirmation that attrition occurred at random, and if not, weighting in analyses was used to account for selective attrition or attrition was very minimal, i.e. no more than 5% (4). Studies in which attrition is likely to have had a substantial influence on study results were considered higher in risk of bias. Too few studies had ≤5% attrition, therefore we deviated from planned analyses and conducted meta-regression to examine the association between attrition and study outcomes.

**4)** Did the study have a smaller sample size? Sampling error is problematic when examining prevalence estimates in small sample sizes. Because sampling error tends to be minimal at sample sizes of ≥1000, studies that were <1000 were considered smaller in sample size and higher in risk of bias.

**5)** Was the pre-pandemic measure of mental health symptomology collected more than 12 months prior to the post-pandemic measure? Studies with measurement of pre-vs.-post mental health symptomology that are separated by a larger period of time may be confounded by other factors (e.g. developmental and age differences in mental health symptomology), therefore we considered studies with a >12 month gap as being higher in risk of bias.

**6)** Was the survey delivery mode (e.g. online) consistent across pre and post outbreak waves of data collection? Because how surveys are administered may affect responses and who takes part, we deemed a change in survey delivery mode (e.g. going from paper-based or face to face interviews pre-pandemic to online post-pandemic outbreak) as increasing risk of bias. This risk of bias indicator was not outlined in our pre-registered protocol and included during eligibility assessment and prior to data extraction and analysis.

**7)** Were conflicts of interests reported? Studies that provided no conflicts of interest statement or report relevant conflicts of interest were considered higher in risk of bias.

#### Influential case and publication bias planned analyses

We characterised outliers as: any effect sizes for which the upper bound of the 95% confidence interval is lower than the lower bound of the pooled effect confidence interval (i.e., extremely small effects) or for which the lower bound of the 95% confidence interval is higher than the upper bound of the pooled effect confidence interval (i.e., extremely large effects). If any outliers are identified we planned to also report the results of meta-analyses with the outliers removed. To address influential cases, we computed DFBETAS values for each effect size. Influential cases were identified as DFBETAS values > 1 (indicative of a >1 change in the standard deviation of the estimated co-efficient after removal of the study (5).

To increase sensitivity, we conducted leave-one-out analyses by removing each study (k) from the primary analyses, and refitting the model. If the removal of k substantially influenced the model (statistical significance of the model changes from $p < .05$ to $p > .05$ (or $p > .05$ to $p < .05$), this was classed as an influential case. We examined evidence for publication bias in our main analyses by examining asymmetry of the effect sizes. We plotted and visually inspected funnel plots for potential publication bias. Next, we conducted an Egger's test of asymmetry and Trim and Fill procedure. For Egger's test, if the intercept is significantly different from 0 at $p > .10$ this is indicative of bias (6). Trim and Fill procedure removes less precise studies which might cause any asymmetry ('trim'), re-estimates the overall effect size, and then replaces removed studies and missing counterparts ('fill') required for symmetry (7). We planned to report i) the number of missing ('filled') studies, and ii) the estimate of the effect size following their inclusion.

National COVID-19 data

*Oxford COVID-19 Government Response Tracker* (OxCGRT)

For each eligible study we extracted the level of severity of social restrictions (e.g. stay at home orders), number of health measures (e.g. contact tracing) and level of economic support (e.g. income support schemes) in place during the month post-pandemic outbreak mental health was assessed as measured by the OxCGRT. (8). OxCGRT systematically collects information on policy responses and interventions that national governments have taken during the pandemic. For each index, governments are scored on an ordinal scale gauging the extent to which a common policy or intervention has been implemented and a sum score is produced, whereby higher scores indicate the extent of the governmental response in a given policy area. Composite indices for each area of policy response are produced for each day of the pandemic for nearly all countries in the world. For each study included in the meta-analysis we examine the governmental response to the pandemic in that country by taking an average of the OxCGRT composite indicators for all days in the month(s) of the pandemic for which mental health data was recorded. For the United States we aggregated OxCGRT composite indicators from the state-level dataset to produce an overall account of the pandemic response in the U.S. for each month examined. In OxCGRT, level of economic support (*Economic Support Index*) is scored on two criteria; presence of income support policies (i.e. to what extent is the government supporting income or providing cash payments to people unable to work), and presence of debt / contract relief policies (i.e. to what extent is the government freezing financial obligations). Severity of social restrictions (*Stringency index*) was assessed across 9 areas gauging the; use of school closures, workplace closures, cancellation of public events, restrictions on gatherings, closure of public transport, use of stay-at-home requirements, restrictions on internal travel, international travel controls, and use of public health information campaigns. A higher total score on the 'Stringency Index' indicates more severe social restrictions. We computed a measure of comprehensiveness of health policies by taking an average of OxCGRT health response components; testing policies, facial covering policies, and vaccination policy in place, whereby a higher total score indicated a great number of COVID-19 health policies in place. For full information on OxCGRT data collection and scoring of individual components, see https://www.bsg.ox.ac.uk/sites/default/files/2020-12/BSG-WP-2020-032-v10.pdf

*Number of cases and deaths*

For each eligible study we used the OxCGRT (9) to extract the number of recorded cases and deaths (by country) for the month that post-pandemic outbreak mental health was measured. Data on COVID-19 attributed deaths in the OxCGRT is collected from *the COVID-19 Data Repository by the Center for Systems Science and Engineering (CSSE)* at Johns Hopkins

University. To account for differences in country population size, we calculated the number of cases and deaths per million population for the month(s) in which post-pandemic outbreak mental health measures were collected in each country.

Meta-analyses methodology

For our main analysis on continuous data, we computed standardised mean change (SMC) in pooled SD units, using the 'escalc:SMCC' function in the R 'metafor' package (9). For analyses examining questionnaire cut-off data we computed Marginal Odds Ratios to quantify size of change ('escalc:MPOR' function). As studies contributed multiple comparisons to analyses, we conducted a multi-level meta-analysis (using the 'rma.mv' function of metafor) to account for heterogeneity within and across studies. We used restricted maximum likelihood models. We planned to determine whether a multi-level meta-analysis was a better fit of the data by a significant loglikelihood ratio test (when compared to a model with study held constant, i.e. a single level model). To calculate standard mean change for our primary analyses we required the correlation between the pre and post pandemic outbreak measure of mental health and we were able to obtain this for 131/177 (74%) of included comparisons. In instances in which data was missing and authors did not respond, we imputed missing correlations based on the average of the obtained correlations (r ~.54). For continuous studies, if the raw data was not available but an effect size was reported (e.g. SMD/C or Odds ratio, we converted if necessary and included it in our analyses). For analyses examining questionnaire cut-offs we computed marginal odds ratios to quantify size of change. Marginal odds ratios were calculated based on the number of individuals who did not meet the questionnaire cut off pre and post pandemic outbreak, number who met the cut off post-pandemic outbreak but not pre-pandemic, number who met the cut-off pre-pandemic outbreak but not post, and the number of individuals who met the cut off at both pre-and post-pandemic outbreak. One study reported a 0 frequency in a cell (Chiaravaolloti), therefore we added 0.5 to each cell in line with the Haldane correction(10).

Use of multi-level vs. single-level meta-analysis in primary analyses

The multi-level model demonstrated that 33.7% of the heterogeneity in effect sizes was attributed to studies and was a better fit of the data than a single level meta-analysis (AIC multi-level model = 30.4, AIC single level model = 82.5: loglikelihood ratio test = 54.09, $p < .001$). Note: results from a single level analysis produced similar results to the main analyses [SMC = .086 [95% CI: .041 to .131], $z = 3.72$, $p < .001$). We retained use of multi-level model analyses for subsequent analyses unless stated otherwise.

Publication bias and influential cases in main analyses

There were no influential cases (DFBETAs ranged -.216 to .216, Cooks' distance < .045). Leave-one-out analysis did not substantially influence the model (ps < .003). Egger's test was significant, suggesting some funnel plot asymmetry ($z = 2.13$, $p = .032$). See figure S1.

Meta-regressions examining attrition and country level predictors of SMC from primary analyses

There was no significant association between level of attrition and change in mental health symptoms characterised by SMC (coefficient = .092 [95% CI: -.185 to .370] $z = .653$, $p = .514$). No country level predictors were significantly associated with SMC; Cases per month ($p = .265$), cases per million ($p = .525$), deaths per month ($p = .831$), deaths per million (p =

.510), Stringency Index (p = .211), level of economic support (p = .797) or country health response index (p = .402).

Figure S1: A funnel plot of all effect sizes in primary meta-analysis.

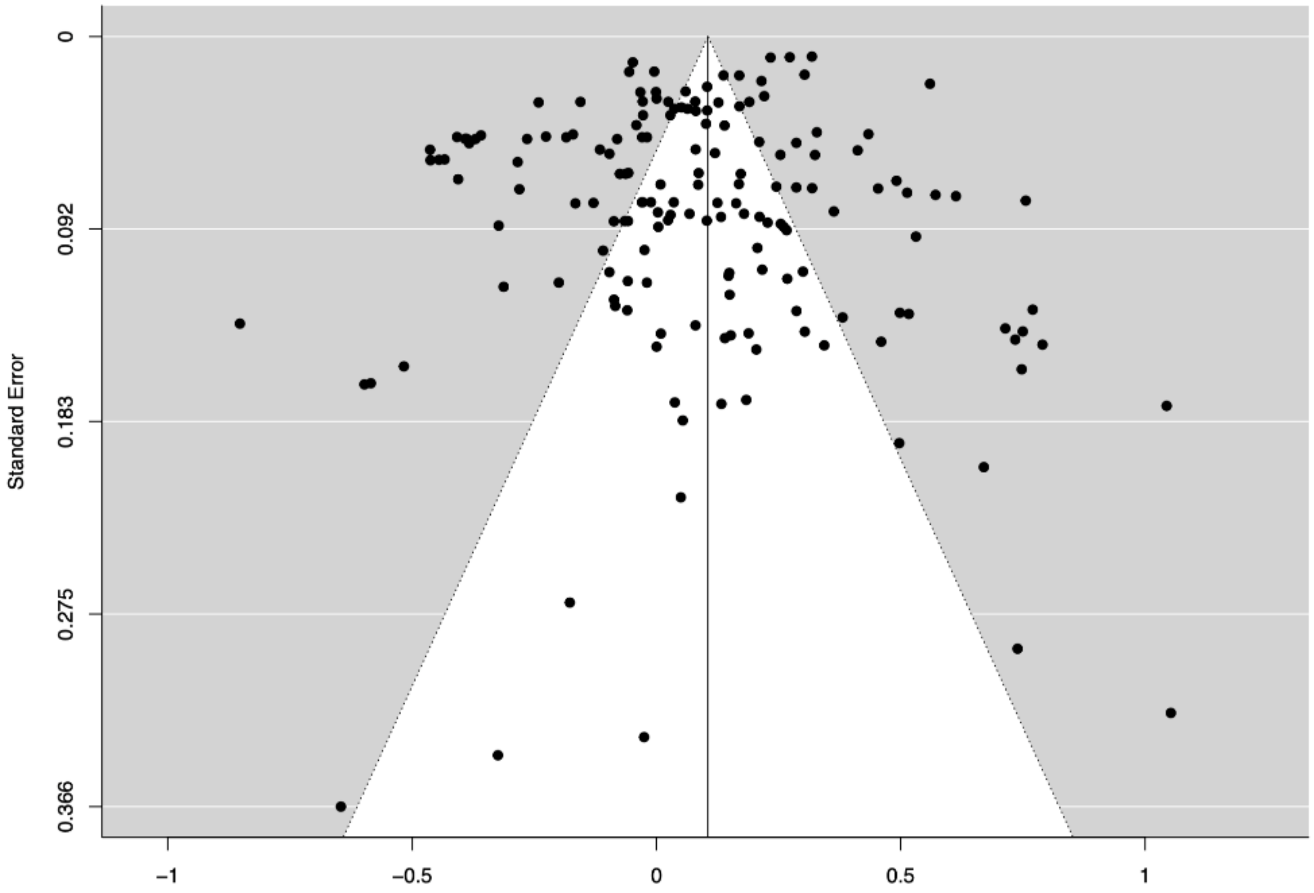

Figure S2. Forest plot of symptom level comparisons: anxiety disorder symptoms

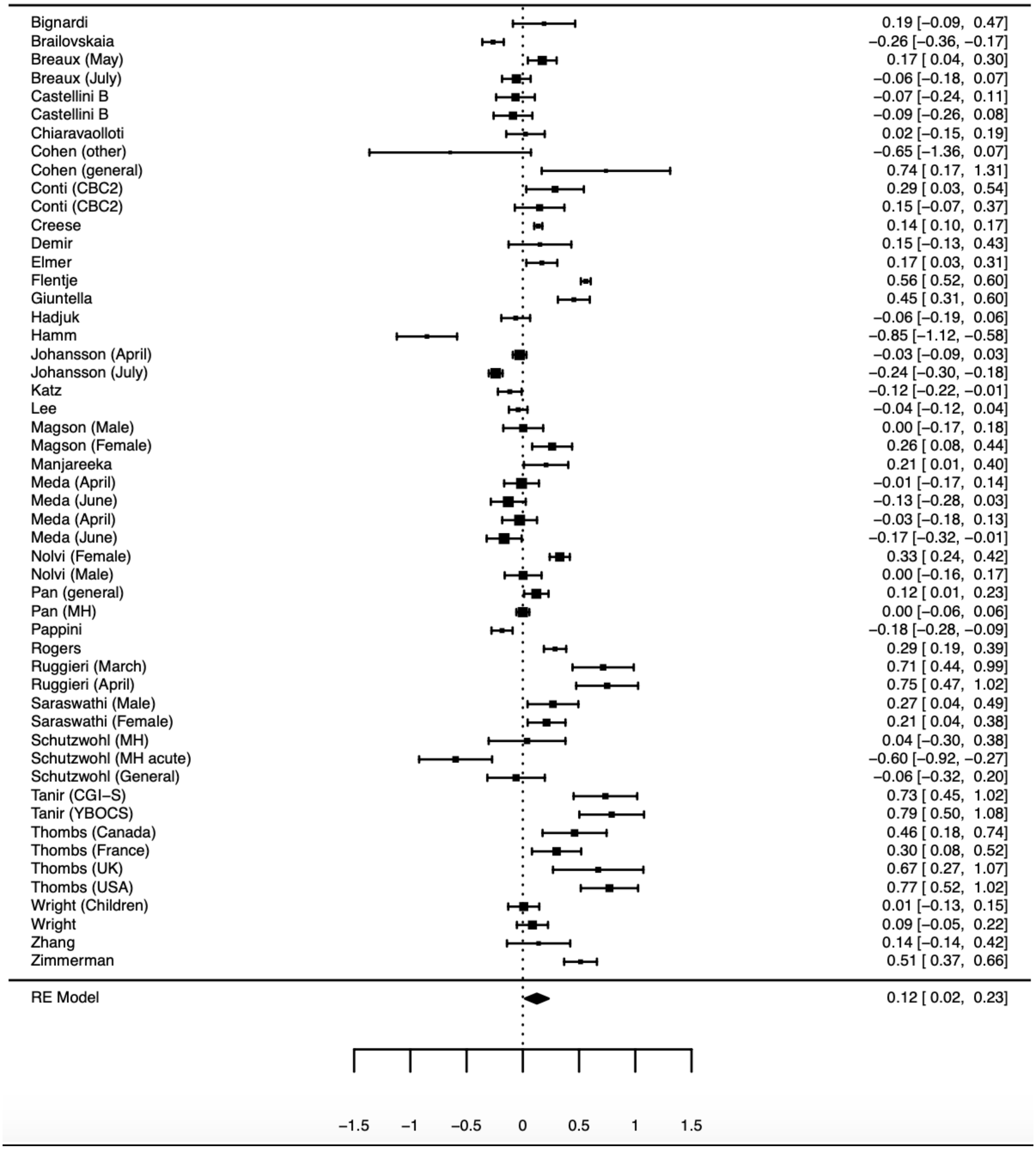


General (general population sample), MH (sample with pre-existing mental health condition), Other (sample from ‘other category’), CBC (Child behaviour checklist), CGI-S (Clinical Global Impression Scale), YBOCS (Children’s Yale-Brown Obsessive Compulsive Scale)

Figure S3. Forest plot of symptom level comparisons: depression and mood disorder symptoms

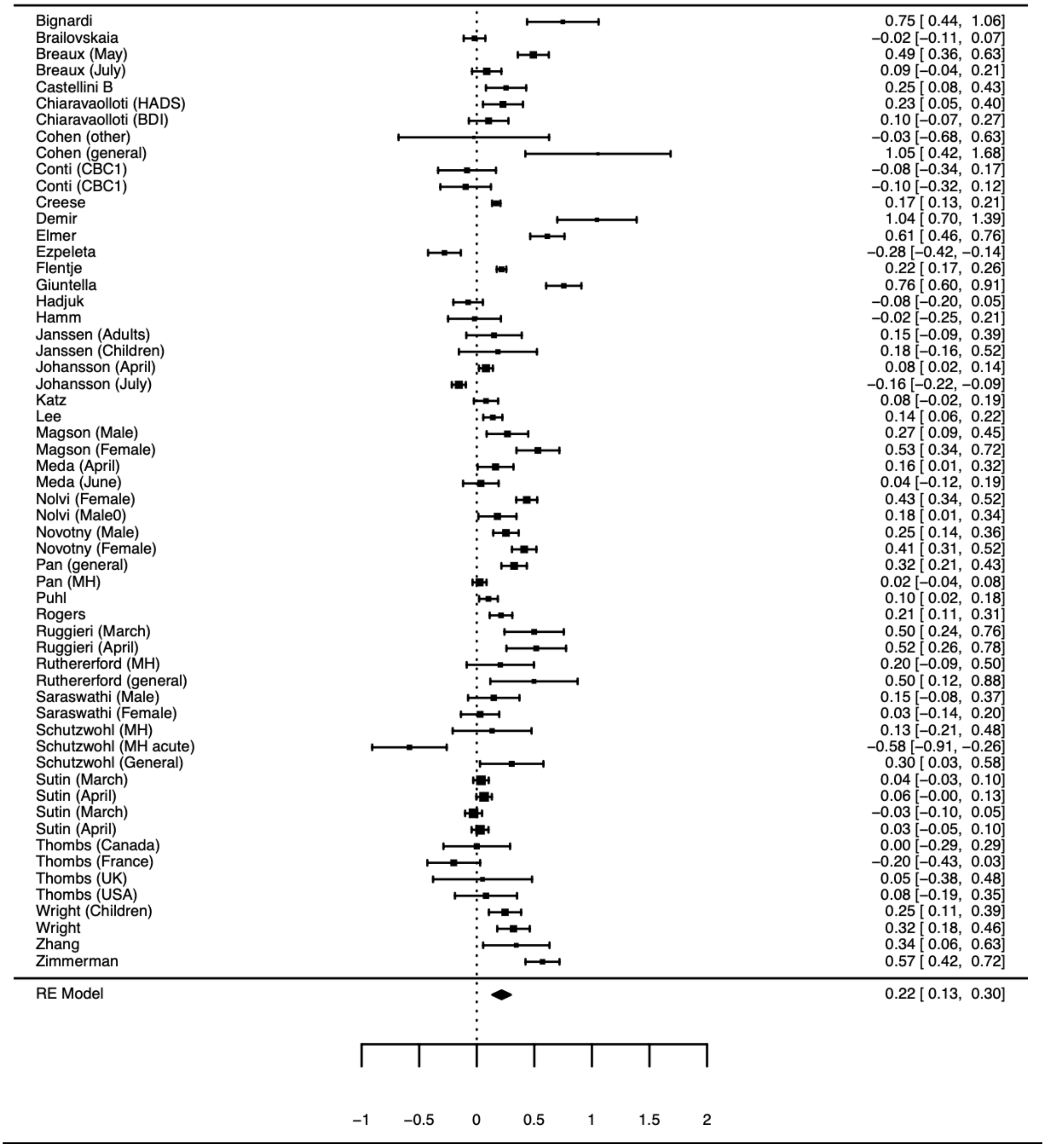


HADS (Hospital Anxiety and Depression scale), BDI (Beck Depression Inventory), General (general population sample), MH (sample with pre-existing mental health condition), Other (sample from ‘other category’), CBC (Child behaviour checklist)

Figure S4. Forest plot of symptom level comparisons: general mental health measures (including distress)

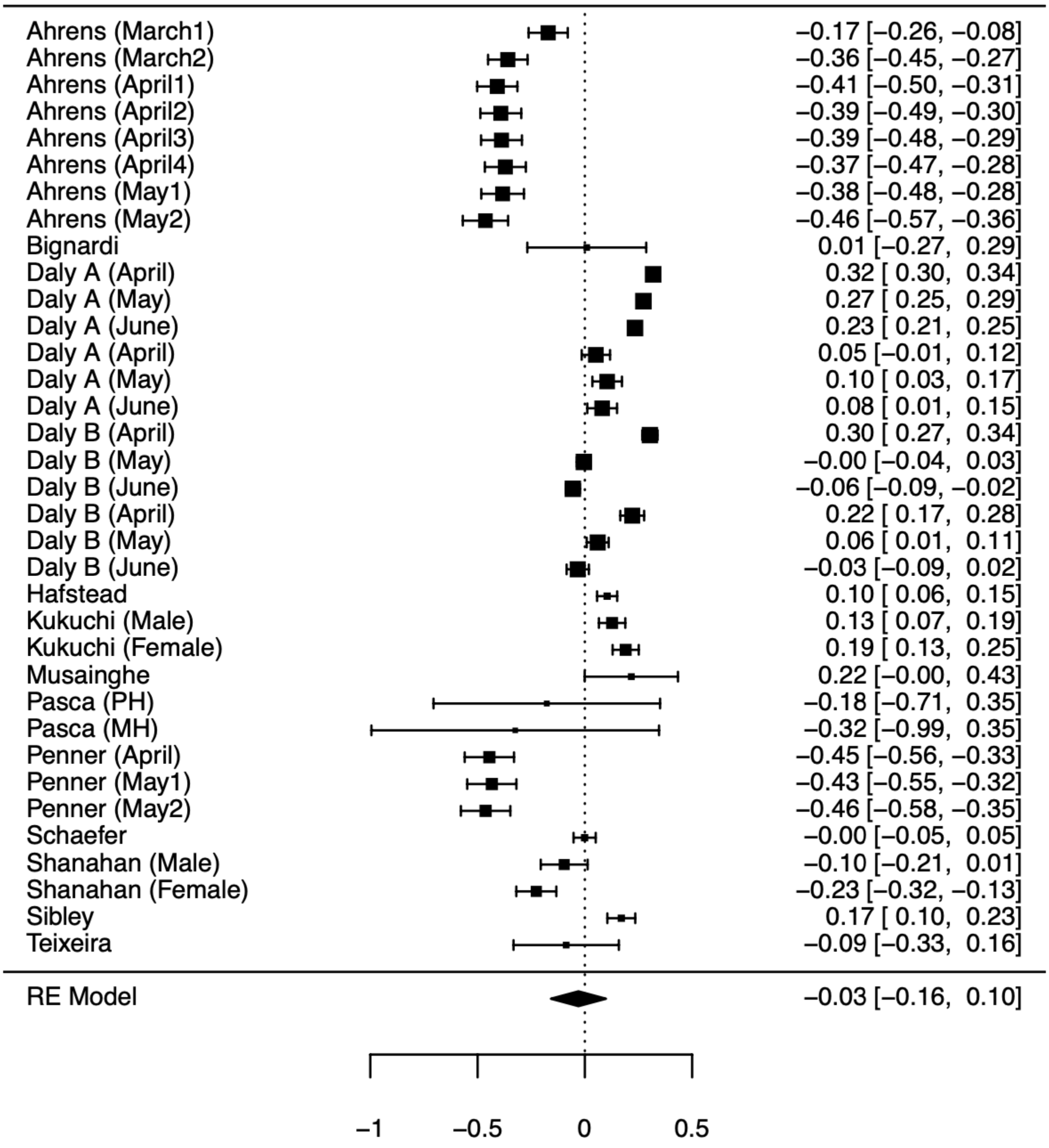

Figure S5. Forest plot of symptom level comparisons: well-being

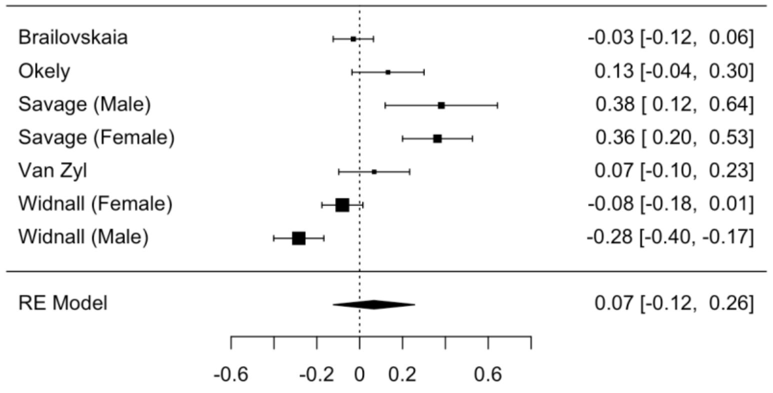


Figure S6. Forest plot of symptom level comparisons: psychotic symptoms

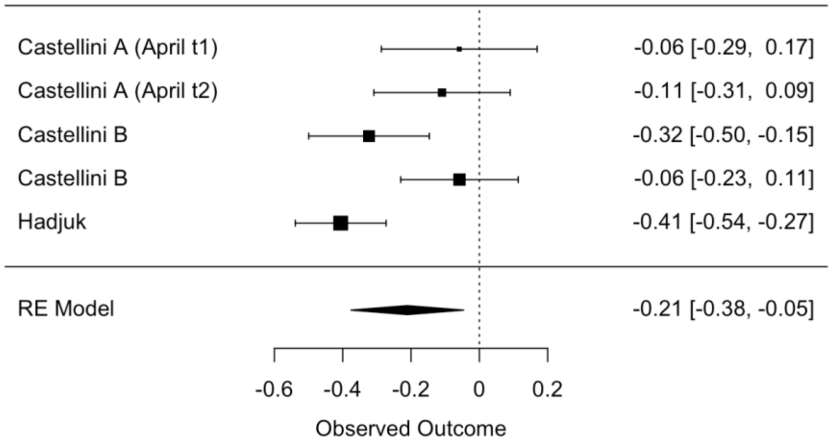


Figure S7. Forest plot of symptom level comparisons: other mental health symptoms

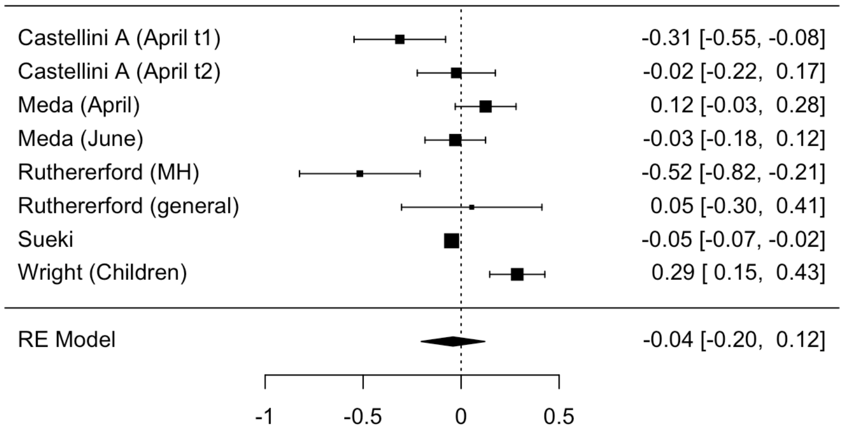


Other symptoms were eating disorder symptoms (Castellini, Meda), PTSD (Rutherford, Wright) and Suicide ideation (Sueki)

Change in numbers exceeding questionnaire cut-offs for mental health problems analysis

A total of 24 comparisons across 12 studies were included in meta-analysis. Sample sizes ranged from 50-7445. Comparisons examined depression and mood disorder symptoms (N=9), anxiety (N=7), general mental-health (N=4), distress (N=2), well-being (N=2). Most comparisons were of adult samples (N=19), as opposed to children/adolescent samples (N=5). The majority of samples were from the general population (N=16), followed by university students (N=6), pre-existing physical health conditions (N=2) and one community sample of Latinx sexual minority men and transgender women. Ten of the 24 comparisons were from March-April 2020 and the remaining 14 comparisons were from May-June. The majority of comparisons were from European samples (N=14), with the remaining samples being mixed (N=2), US (N=1), India (N=5) and Japan (N=2). Attrition ranged from 5%-83%. It was rare for studies to have a consistent mode of delivery (N = 4, 17%) or adopt representative sampling (N=7, 29%). Multilevel meta-analysis was not a better fit of the data than a single level meta-analysis (AIC single level = 31.04, AIC multilevel = 31.72: loglikelihood ratio test = 1.31, p = .252). There was a significant effect (single level meta-analysis), with the odds of exceeding a questionnaire cut-off for mental health problems from pre-post pandemic = 1.31 (Marginal Odds Ratio = 1.31 [95% CI: 1.09 to 1.55], z = 3.18, p = .001, I2 = 93.2%). Trim and fill analysis did not impute any effect sizes. All models were significant in leave one out analysis, with the smallest pooled Marginal Odds Ratio = 1.27, and largest = 1.36. There was no difference in effect sizes between anxiety and depression symptoms (QM(1) = 0.259, p = .611). Three comparisons examined change in mental health symptoms among Chinese samples but due to there being too few comparisons for meaningful analysis, we did not meta-analyse these comparisons.

Table S1. Individual study information for all included articles

| Author | Country | Participant group | Pre pandemic wave | Post pandemic outbreak waves | Mental health measure(s) | Change data | Sample size |
|---|---|---|---|---|---|---|---|
| Ahrens(11) | Germany | Adults in a cohort study | December 2019 - March 2020 | March 23 – May 11 (8 assessments) | GHQ-28 (Distress/non-specific mental health) | Continuous | N=382-482 |
| Bignardi (12) | UK | Children in a cohort study | June 2018 - September 2019 | April 4 – June 6 | SDQ emotional problems, RCADS depression (Depression & mood disord) | Continuous | N=50-51 |
| | | | | | RCADS anxiety (Anxiety disord & symp) | | |
| Brailovskaia(13) | Germany | University sample | October 2019 | March 20-28 | DASS21 depression (Depression & mood disord) | Continuous | N=436 |
| | | | | | DASS21 Anxiety (Anxiety disord & symp) | | |
| | | | | | Unidimensional positive MH scale (Well-being) | | |
| Breaux(14) | US | Adolescents with and without ADHD | September 2018 – February 2020 | May 15 – August 5 (2 assessments) | RCADS depression (Depression & mood disord) | Continuous | N=238 |
| | | | | | RCADS anxiety (Anxiety disord & symp) | | |
| Castellini (A)(15) | Italy | Eating disorder outpatients and healthy controls | November 2019 – January 2020 | April 22 – May 3 | BSI total (Psychotic symptoms) | Continuous | N=74-97 |
| | | | | | Eating disorders examination (Other MH conditions) | | |
| Castellini (B)(16) | Italy | University sample | December 1 2019 – January 2020 | 22 April – May 3 | BSI depression (Depression & mood disord) | Continuous | N=130 |
| | | | | | BSI anxiety, OCD (Anxiety disord & symp) | | |
| | | | | | BSI paranoia, psychoticism (Psychotic symp) | | |
| Chen(17) | China (Hong Kong) | School children | October – November 2019 | March 4 – 16[t] | DASS depression (Depression & mood disord) | Continuous | N=543 |
| | | | | | DASS anxiety (Anxiety disord & symp) | | |

| | | | | | | | |
|---|---|---|---|---|---|---|---|
| Chiaravalloti(18) | US, Canada, Belgium, Denmark, Italy, UK | Multiple sclerosis patients | August – October 2019 | May 4 – July 15 | HADS depression, Beck Depression Inventory II (Depression & mood disord)<br>HADS anxiety (Anxiety disord & symp) | Continuous & Cut-off | N=131 |
| Cohen(19) | US | Adolescents with/without early life stress | July 2019 – February 2020 | May 22 – June 18 | PROMIS depression (Depression & mood disord)<br>PROMIS anxiety (Anxiety disord & symp) | Continuous | N=9-15 |
| Conti(20) | Italy | Patients with neurological or psychiatric disorders | September 2019 – February 2020 | April 20 – May 4 | CBC affective (Depression & mood disord)<br>CBC anxiety (Anxiety disord & symp) | Continuous | N=61-80 |
| Creese(21) | UK | Adults aged 50 and over in a cohort study | 2015-2019 | May 13 – June 8 | PHQ9 (Depression & mood disord)<br>GAD7 (Anxiety disord & symp) | Continuous | N=2,959 |
| Daly (A)(22) | UK | Adults in a cohort study | 2017-2019 | April 24 – July 1 (X3assessments) | GHQ12 (Distress/non-specific mental health) | Continuous | N=801–11,599 |
| Daly (B)(23) | US | Adults in a cohort study | March 2020 | April 1 – June 23<br>(3 assessments) | PHQ4 (Distress/non-specific mental health) | Continuous | N=1,275-3,609 |
| Demir(24) | Turkey | MS patients | January 2020 | April 23 | Beck inventory (Depression & mood disord)<br>Beck inventory (Anxiety disord & symp) | Continuous | N=50 |
| Dickerson(25) | UK | Community sample | June 2017 – March 2020 | April 10 – June 30 | PHQ8 (Depression & mood disord)<br>GAD7 (Anxiety disord & symp) | Cut-off | N=1,634-1,730 |
| Elmer(26) | Switzerland | University sample | September 2019 | April | CES-D (Depression & mood disord)<br>GAD (Anxiety disord & symp) | Continuous | N=206 |

| | | | | | | | |
|---|---|---|---|---|---|---|---|
| Ezpeleta(27) | Spain | Families in a community sample | Unspecified (2019) | June | SDQ emotional problems (Depression & mood disord) | Continuous | N=197 |
| Flentje(28) | US | Sexual and gender minority cohort with and without existing MH condition | June 2019 | March 23 – April 19 | PHQ9 (Depression & mood disord)<br>GAD7 (Anxiety disord & symp) | Continuous | N=671-1610 |
| Giuntella(29) | US | University sample | Spring 2019 | March 20 | CES-D (Depression & mood disord)<br>GAD7 (Anxiety disord & symp) | Continuous | N=211 |
| Groot(30) | Denmark | Adults in a cohort study | March 2016 – February 2020 | March 27-April | Short WEMBWS (Well-being) | Cut-off | N=7,445 |
| Hafstead(31) | Norway | Adolescents in a cohort study | February 2019 | June | Hopkins symptom checklist (Distress/non-specific mental health) | Continuous & Cut-off | N=1,763 |
| Hajduk(32) | Slovakia | University sample | October 2018 | April | PHQ9 (Depression & mood disord)<br>GAD7 (Anxiety disord & symp)<br>Community assessment of psychic experiences (Psychotic symp) | Continuous | N=235 |
| Hamm(33) | US | Older adults with depression | Unspecified, most participants 2019 | April 1 – April 23 | PHQ9 (Depression & mood disord)<br>PROMIS anxiety (Anxiety disord & symp) | Continuous | N=73 |
| Janssen(34) | Netherlands | Parents of adolescents and adolescents from cohort | September 2018 – November 2019 | April 14 – April 28 | PHQ9 (Depression & mood disord) | Continuous | N=34-67 |
| Johansson(35) | Sweden | University sample | August 2019-March 2020 | March 14 – 10 September (2 assessments) | DASS depression (Depression & mood disord)<br>DASS anxiety (Anxiety disord & symp) | Continuous | N=1,049 |

| | | | | | | | |
|---|---|---|---|---|---|---|---|
| Katz(36) | US, Canada, UK and Ireland | Adults from an online panel | April 2019 | April 15-20 | DASS depression (Depression & mood disord)<br>DASS anxiety (Anxiety disord & symp) | Continuous | N=348 |
| Kikuchi(37) | Japan | Adults from an online panel | February 2020 | April 1 - 6 | Kessler 6 (Distress/non-specific mental health) | Continuous & Cut-off | N=1,024-1,054 |
| Kwong(38) | UK | Young adults in a cohort study | 2017-2018 | April 9 – May 14 | SMFQ (Depression & mood disord)<br>GAD7 (Anxiety disord & symp)<br>Short WEMWBS (Well-being) | Continuous & Cut-off | N=2,812-2,850 |
| Lee(39) | US | Community sample of young adults | January 2020 | April | PHQ4 (Depression & mood disord)<br>PHQ4 (Anxiety disord & symp) | Continuous | N=564 |
| Linnemayr(40) | US | Community sample of Latinx sexual minority men and transgender women | April 2018-July 2019 | May 1 - May 18 | Kessler (Distress/non-specific mental health) | Cut-off | N=50 |
| Magson(41) | Australia | Community sample of adolescents | 2019 | May 5 – May 14 | SMFQ (Depression & mood disord)<br>SCAS GAD (Anxiety disord & symp) | Continuous | N=122-126 |
| Manjareeka(42) | India | University sample (medical students) | February 2020 | May | State-trait anxiety scale (Anxiety disord & symp) | Continuous & Cut-off | N=101 |
| Meda(43) | Italy | University students | October – December 2019 | April 3 – June 21 | Beck depression inventory (Depression & mood disord) | Continuous | N=161-197 |

| | | | | | | | |
|---|---|---|---|---|---|---|---|
| | | | | | Beck anxiety inventory, OC inventory revised (Anxiety disord & symp)<br>Eating disorder inventory (Other MH conditions) | | |
| Munasinghe(44) | Australia | Adolescent community sample | November 2019 – March 2020 | March 23 – April 19 | Kessler 6 (Distress/non-specific mental health) | Continuous | N=421 |
| Nolvi(45) | Finland | Parents in birth cohort study | 2014-2019 | May 4 – June 7 | Edinburgh postnatal depression scale (Depression & mood disord)<br>Symptom checklist 90 (Anxiety disord & symp) | Continuous & Cut-off | N=143-508 |
| Novotny(46) | Czech Republic | Adults in a cohort study | 2015 | April 24 – May 27 | PHQ (Depression & mood disord) | Continuous | N=326-370 |
| Okely(47) | Scotland | Older adults in cohort study | 2017-2019 | May 27 | Short WEMBWS (Well-being) | Continuous | N=137 |
| Pan(48) | Netherlands | People with/ without MH condition in cohort studies | 2006-2016 | April 1 – May 13 | QIDS (Depression & mood disord)<br>Beck anxiety inventory (Anxiety disord & symp) | Continuous | N=328-1148 |
| Pappini(49) | US | University sample | January 2020 | March 27-May | GAD7 (Anxiety disord & symp) | Continuous | N=443 |
| Pasca(50) | Italy | Adolescents with epilepsy & neurocognitive or internalising disorders | September 2019-Feburary 2020 | April – May | CBC internalizing (Distress/non-specific mental health) | Continuous | N=9-14 |
| Penner(51) | US | Community sample of Hispanic/Latinx adolescents | January 2020 | April – May (3 assessments) | Brief Problem Monitor- internalizing symps (Distress/non-specific mental health) | Continuous | N=320 |

| | | | | | | | |
|---|---|---|---|---|---|---|---|
| Puhl(52) | US | Secondary school students | 2017-2018 | April - June | Kandal and Davies depression scale (Depression & mood disord) | Continuous | N=584 |
| Rogers(53) | US | Adults in cohort study | October 2019 | April 11 - 15 | CDI (Depression & mood disord)<br>GAD (Anxiety disord & symp) | Continuous | N=407 |
| Ruggieri(54) | Italy | Adult sample of social media users | March 2020 | March 25 – April 14 (2 assessments) | DASS21 depression (Depression & mood disord)<br>DASS21 anxiety (Anxiety disord & symp) | Continuous | N=65 |
| Rutherford(55) | US | Older adults diagnosed with PTSD and trauma exposed healthy controls | Unspecified, prior to March 2020 | April 1 – May 8 | PTSD checklist (Other MH conditions)<br>Hamilton scale (Depression & mood disord) | Continuous | N=30-46 |
| Saraswathi(56) | India | University sample (medical students) | December 2019 | June 10-20 | DASS21 depression (Depression & mood disord)<br>DASS21 anxiety (Anxiety disord & symp) | Continuous & Cut-off | N=78-139 |
| Savage(57) | UK | University sample | October 2019 | April | WEMWBS (Well-being) | Continuous | N=60-154 |
| Schaefer(58) | Germany | Community sample of adults | February 2020 | March 16-20 | Mini-symptom checklist total (Psychotic symp) | Continuous | N=1439 |
| Schützwohl(59) | Germany | Adults with mental health conditions and no conditions | August 2019 – March 2020 | April 22 – May 13 | BSI-18 depression (Depression & mood disord)<br>BSI-18 anxiety (Anxiety disord & symp) | Continuous | N=33-59 |
| Shanahan(60) | Switzerland | Adults in community sample | April – September 2018 | April 11-18 | Social Behaviour Questionnaire Internalising (Distress/non-specific mental health) | Continuous | N=323-453 |
| Shan Wong(61) | China (Hong Kong) | Older adults in primary care | April 2018 – March 2019 | March 24 – April 15 | PHQ9 (Depression & mood disord) | Continuous & Cut-off | N=583 |

| | | | | | | | |
|---|---|---|---|---|---|---|---|
| | | | | | GAD7 (Anxiety disord & symp) | | |
| Sibley(62) | New Zealand | Adults in cohort study | October – December 2019 | March 26 – April 12 | Kessler 6 (Distress/non-specific mental health) | Continuous | N=921 |
| Sueki(63) | Japan | Adults from an online panel | January 2020 | April 27-30 | Suicidal ideation scale (Other MH conditions) | Continuous | N=6,683 |
| Sutin(64) | US | Adults from an online panel | January – February 2020 | March 18 – April 29 (2 assessments) | PHQ2 (Depression & mood disord) | Continuous | N=719-858 |
| Tanir(65) | Turkey | Children and adolescents with OCD | September 2019 – March 2020 | April 20 - 30 | Clinical Global Impression Scale, Children's Yale-Brown Obsessive Compulsive Scale (Anxiety disord & symp) | Continuous | N=61 |
| Teixeira(66) | Brazil | Older women enrolled in an exercise programme | January – February 2020 | June | World Health Organisation quality of life mental health subscale (Distress/non-specific mental health) | Continuous | N=64 |
| Thombs(67) | US, Canada, UK, France | Patients with multiple sclerosis | July – December 2019 | April 9 - 27 | PHQ8 (Depression & mood disord)<br>PROMIS anxiety (Anxiety disord & symp) | Continuous | N=43-159 |
| Van der Velden(68) | Netherlands | Adults from an online panel | November 2019 | June | Mental health inventory (Distress/non-specific mental health) | Cut off | N=4,084 |
| Van Zyl(69) | Netherlands | University sample | January 2020 | April | Mental health continuum short-form (Well-being) | Continuous | N=141 |
| Wen Li(70) | China | University sample | November 2019 | February – June (2 assessments) | DASS21 (Depression & mood disord)<br>DASS21 (Anxiety disord & symp) | Continuous | N=173 |
| Widnall(71) | UK | High school students | October 2019 | April - May | HADS anxiety (Anxiety disord & symp)<br>HADS depression (Depression & mood disord)<br>WEMBWS (Well-being) | Continuous & Cut-off | N=292-458 |

| | | | | | | | |
|---|---|---|---|---|---|---|---|
| Wright(72) | UK | Adolescents and parents in a birth cohort | December 2019 – March 2020 | June 18 – August 4 | SMFQ, PHQ9 (Depression & mood disord)<br>Child trauma symptom scale (Other MH conditions)<br>Spence anxiety scale, GAD7 (Anxiety disord & symp) | Continuous | N=202 |
| Yan Li(73) | China | University sample | December 2019 | February 4 - 6 | PHQ4 (Distress/non-specific mental health) | Continuous | N=555 |
| Zhang(74) | US | University sample | January 2020 | May | PHQ9 (Depression & mood disord)<br>GAD7 (Anxiety disord & symp) | Continuous | N=49 |
| Zimmerman(75) | US | University sample | February 2020 | April 3 - 20 | PHQ9 (Depression & mood disord)<br>GAD7 (Anxiety disord & symp) | Continuous | N=205 |

BSI (Brief symptom inventory), CBC (Child behaviour checklist), CDI (Children’s depression inventory), DASS (Depression, anxiety and stress scale), GAD (Generalised anxiety disorder scale), GHQ (General health questionnaire), HADS (Hospital anxiety and depression scale), PHQ (Patient health questionnaire), PROMIS (Patient-Reported Outcomes Measurement Information system), QIDS (Quick Inventory of Depressive Symptomatology), RCADS (Revised children’s anxiety and depression scale), SCAS (Spence Children's Anxiety Scale), SDQ (Strengths and difficulties questionnaire), SMFQ (Short mood and feelings questionnaire), WEMWBS (Warwick-Edinburgh mental well-being scale).

Table S2.Risk of bias ratings for individual studies

| Author | Publication status | Representative sampling | Consistency of survey delivery mode | Attrition | Small sample size (N<1000) | More than 12 months between pre and post pandemic measures | Study reports no relevant conflicts of interest |
|---|---|---|---|---|---|---|---|
| Ahrens | J | N/U | N/U | U | Y | N | N/U |
| Bignardi | PP | Y | Y | 92% | Y | Y | Y |
| Brailovskaia | J | N/U | Y | 13% | Y | N | N/U |
| Breaux | J | N/U | N/U | 9% | Y | Y | N/U |
| Chiaravalloti | J | N/U | N/U | 5% | Y | N | N/U |
| Castellini (A) | J | N/U | N/U | 3-10% | Y | N | Y |
| Castellini (B) | J | N/U | Y | 15% | Y | N | Y |
| Chen | J | N/U | N/U | 51% | Y | N | Y |
| Cohen | PP | N/U | N/U | U | Y | N | N/U |
| Conti | J | N/U | Y | U | Y | N | Y |
| Creese | J | N/U | Y | U | N | Y | Y |
| Daly (A) | J | Y | N/U | 62-66% | Mixed | Y | Y |
| Daly (B) | J | Y | Y | 11-21% | N | N | Y |
| Demir | J | N/U | Y | U | Y | N | Y |
| Dickerson | PP | N/U | N/U | U | N | Y | Y |
| Elmer | J | N/U | Y | U | Y | N | Y |
| Ezpeleta | J | N/U | N/U | 52% | Y | Y | Y |
| Flentje | J | N/U | N/U | N/U | Mixed | N | N/U |
| Giuntella | PP | N/U | Y | U | Y | N | N/U |
| Groot | pp | N/U | N/U | 83% | N | Y | Y |
| Hafstead | PP | Y | N/U | 83% | N | Y | Y |
| Hajduk | J | N/U | N/U | 82% | Y | Y | Y |
| Hamm | J | N/U | N/U | 34% | Y | Y | N/U |
| Janssen | J | N/U | N/U | 56-58% | Y | Y | Y |
| Johansson | PP | N/U | Y | 43% | N | N | Y |
| Katz | PP | N/U | Y | 23% | Y | N | N/U |
| Kikuchi | J | Y | Y | 14% | N | N | Y |
| Kwong | J | Y | N/U | 81% | N | Y | Y |

| | | | | | | | |
|---|---|---|---|---|---|---|---|
| Lee | J | N/U | Y | U | Y | N | Y |
| Linnemayr | J | N/U | N/U | U | Y | Y | Y |
| Magson | J | N/U | Y | 47% | Y | Y | Y |
| Mankareeka | J | N/U | Y | 15% | Y | N | N/U |
| Meda | J | N/U | Y | U | Y | N | Y |
| Munasinghe | J | N/U | Y | 68% | Y | N | Y |
| Nolvi | PP | N/U | N/U | U | Y | Y | Y |
| Novotny | J | Y | N/U | 61% | Y | Y | Y |
| Okely | J | N/U | N/U | 67% | Y | Y | Y |
| Pan | J | N/U | N/U | 42% | Mixed | Y | Y |
| Pappini | PP | N/U | Y | U | Y | N | Y |
| Pasca | J | N/U | N/U | U | Y | N | Y |
| Penner | J | N/U | N/U | 18% | Y | N | Y |
| Puhl | J | N/U | N/U | 63% | Y | Y | Y |
| Rogers | J | Y | Y | 33% | Y | N | Y |
| Ruggieri | J | N/U | Y | 65% | Y | N | Y |
| Rutherford | J | N/U | N/U | U | Y | N/U | Y |
| Saraswathi | PP | N/U | N/U | 9% | Y | N | Y |
| Savage | J | N/U | Y | 86% | Y | N | Y |
| Schaefer | J | N/U | Y | 21% | N | N | Y |
| Schützwohl | PP | Y | N/U | 9-35% | Y | N | Y |
| Shanahan | J | N/U | N/U | 32% | Y | Y | Y |
| Shan Wong | J | N/U | N/U | 22% | Y | Y | Y |
| Sibley | J | Y | N/U | Unclear | Y | N | Y |
| Sueki | PP | Y | Y | 33% | N | N | N/U |
| Sutin | J | Y | Y | 60% | Y | N | Y |
| Tanir | J | N/U | N/U | Unclear | Y | N | Y |
| Teixeria | PP | N/U | N/U | Unclear | Y | N | Y |
| Thombs | J | N/U | Y | 63-67% | Y | N | Y |
| Van der Velden | J | Y | Y | 33% | N | N | Y |
| Van Zyl | PP | N/U | Y | Unclear | Y | N | N/U |
| Wen Li | J | N/U | N/U | 15% | Y | N | Y |
| Widnall | PP | N/U | N/U | 70% | Y | N | N/U |
| Wright | PP | N/U | N/U | 11% | Y | N | Y |

| | | | | | | | |
|---|---|---|---|---|---|---|---|
| Yan Li | J | N/U | N/U | 11% | Y | N | Y |
| Zhang | J | N/U | N/U | 0% | Y | N | Y |
| Zimmerman | PP | N/U | Y | 26% | Y | N | N/U |

J (Journal), PP (pre-print), N (No), U (Unclear), Y (Yes). Mixed (some comparisons from study were smaller than N < 1000 participants and some were larger)
